# The Cost-effectiveness of a Continuous Glucose Monitoring Device for Adult Diabetes Patients in Ethiopia: A Semi-Markov Modelling Study

**DOI:** 10.1101/2025.01.05.25320005

**Authors:** Michael Endale Mengesha

## Abstract

**Background:** Diabetes Mellitus is a chronic condition requiring regular blood glucose monitoring to prevent acute complications and long-term vascular damage. While standard glucometers are commonly used, continuous glucose monitoring (CGM) devices offer real-time insights into glucose fluctuations, improving glycemic control. However, cost-effectiveness data on CGM use in low-resource settings like Ethiopia remains limited.

**Methods:** A semi-Markov model was developed to evaluate the cost-effectiveness of CGM compared to standard self-monitoring of blood glucose in adult diabetes patients in Ethiopia. The analysis incorporated costs from a societal perspective and lifetime horizon, health utilities, and transition probabilities from local and global sources. Primary outcomes included incremental cost-effectiveness ratios (ICERs) and net monetary benefit (NMB) at a willingness-to-pay threshold of one times Ethiopia’s GDP per capita (ETB 165,000).

**Results:** CGM was associated with ICERs of ETB 155,625, ETB 135,353, and ETB 59,718 per QALY gained for males aged 20, 40, and 60 years, respectively. For females, the ICERs were higher at ETB 156,881, ETB 140,290, and ETB 69,791 per QALY for the same age groups, with cost-effectiveness improving with age. Net monetary benefit (NMB) results were positive for the average cohort starting age of 20, 40 and 60 in both male and female categories. One-way and probabilistic sensitivity analyses confirmed the robustness of these findings, with cost and health outcome discount rates identified as key drivers of parameter uncertainty.

**Conclusion:** CGM is a cost-effective intervention for diabetes management in Ethiopia, particularly in older adults. Its adoption could reduce complications and improve health outcomes, but affordability barriers for younger populations warrant further policy considerations.

## Background

Diabetes Mellitus, colloquially diabetes, is a chronic disease that occurs either when the pancreas does not produce enough insulin, a hormone that regulates blood glucose level, or when the body cannot effectively use the insulin it produces (1). According to the International Diabetes Federation the prevalence of diabetes in Ethiopia in 2021 was 3.3% with 1.9 million total adult cases (2). Moreover, studies have shown a concerning growing trend in the prevalence of diabetes in Ethiopia (3,4).

Diabetes patients are advised to frequently monitor the amount of glucose in their blood and administer the appropriate treatment to keep their blood glucose levels in the target range (5). Out of range blood glucose levels, either high or low, may lead to several acute life-threatening conditions such as loss of consciousness, seizure or coma. Over the long term, poorly controlled diabetes leads to the development of diabetes related microvascular (such as kidney disease, retinopathy, and neuropathy) and macrovascular (such as stroke, myocardial infarction, and heart failure) complications (6,7).

Typically, people with diabetes self-monitor their blood glucose levels using a blood glucose meter. In order to do so, a person must prick their finger and squeeze a drop of blood onto a test strip inserted into the meter. The meter device then provides a readout of the blood glucose level. People with diabetes who use a meter usually take readings at regular intervals, including before meals, after meals, before and after physical activity, before driving, and during the night.

### Health Technology Under Review

Continuous glucose monitoring (CGM) provides an opportunity for patients to monitor their blood glucose levels more frequently. It is aimed at helping people with diabetes gain a better understanding of their blood glucose control in real time.

Continuous glucose monitors consist of a sensor inserted underneath the skin, a transmitter, and a small monitor. Every few minutes, the sensor measures blood glucose levels in the interstitial fluid and sends readings via the transmitter to the monitor, which displays the information. For some models, the information can also be transmitted to other devices using near field communication (NFC) and Bluetooth technology (5). These devices also feature systems for directly sharing results with clinicians and linking results to smartphone applications (4). The sensors for most of the currently available continuous glucose monitors are intended to be used for no more than 14 days and must be replaced regularly.

### International Context

The first continuous glucose monitoring system was approved by the FDA in 1999, since then it has garnered widespread use around the world, predominantly in developed nations (5,8). The market for CGM devices is currently dominated by two manufacturers, Abbott Technologies and DexCom, mainly differing in entry price points and customization abilities (9). Recently there have been regulatory approvals of newer non-invasive glucose sensing systems that utilize radiofrequency and spectroscopy techniques (8).

### Ethiopia Context

A situational analysis done on mostly urban settings found that a majority of clinicians and patients have not been previously informed about digital continuous glucose monitoring devices. However, they welcomed and expressed keen interest in utilizing these devices and were open to trying them out (4). It is generally seen as a positive factor, although concerns about technology adoption and affordability are raised.

### Clinical Evidence

There are several clinical and observational studies showing the benefits of continuous glucose monitoring in reducing the time patients spent outside the target glucose range and increasing the time spent within the target range. This time-related glucose variability is considered to be a superior measure to Hemoglobin A1C (HgA1C) since the latter only accounts for 3-month average and might mask the daily swings of blood glucose levels (5).

## Methods

A Markov model with time dependency, hence semi-markov model was constructed to assess the cost-effectiveness of continuous glucose monitoring when compared to the usual care of self-monitoring of blood glucose levels.

### Outcome measures

The following outcomes were examined: incremental costs, incremental quality-adjusted life-years (QALYs), incremental cost-effectiveness ratios (ICERs), net monetary benefit (NMB) and cost effectiveness acceptability curves (CEAC). The WHO’s Choosing Cost-Effective Interventions (CHOICE) describes interventions with an ICER per QALY of less than one times the gross domestic product (GDP) per capita of a country as “very cost-effective,” of one to three times the GDP per capita as “cost-effective,” and of greater than three times the GDP per capita as “not cost-effective.” (10). However, there is no consensus to which threshold value to use for health interventions in Ethiopia. In this paper, a threshold value of one times current GDP per capita in Ethiopia is used in the base case scenario. Different threshold values were tested and reported under sensitivity analysis.

### Target population

The population of interest was adult patients (age ≥ 20 years) with diabetes mellitus who require frequent blood glucose tests and are on insulin therapy.

### Perspective

The analysis was done taking into consideration costs and benefits from a societal perspective and a lifetime horizon. Both the costs and health outcomes were discounted at the rate of 3% in the base case scenario.

### Interventions

Continuous glucose monitoring can be performed using different devices and technologies (see Background). Since clinical effectiveness and market information on these devices is limited, the base case scenario will focus on the widely used and cheaper alternative stand-alone CGM device, Abbott’s FreeStyle Libre 2.

### Model structure and assumptions

Using Excel *version 2411*, a Markov process model for an overtly simplified diabetes disease progression was developed. Four mutually exclusive health states were used in the model (Fig 1). Since the effectiveness of continuous glucose monitoring has been shown in adults, in this model, we followed a hypothetical Ethiopian population cohort from age 20 over their lifetime (i.e, starting with an adult population of diabetes patients without complications).

**Figure 1.**
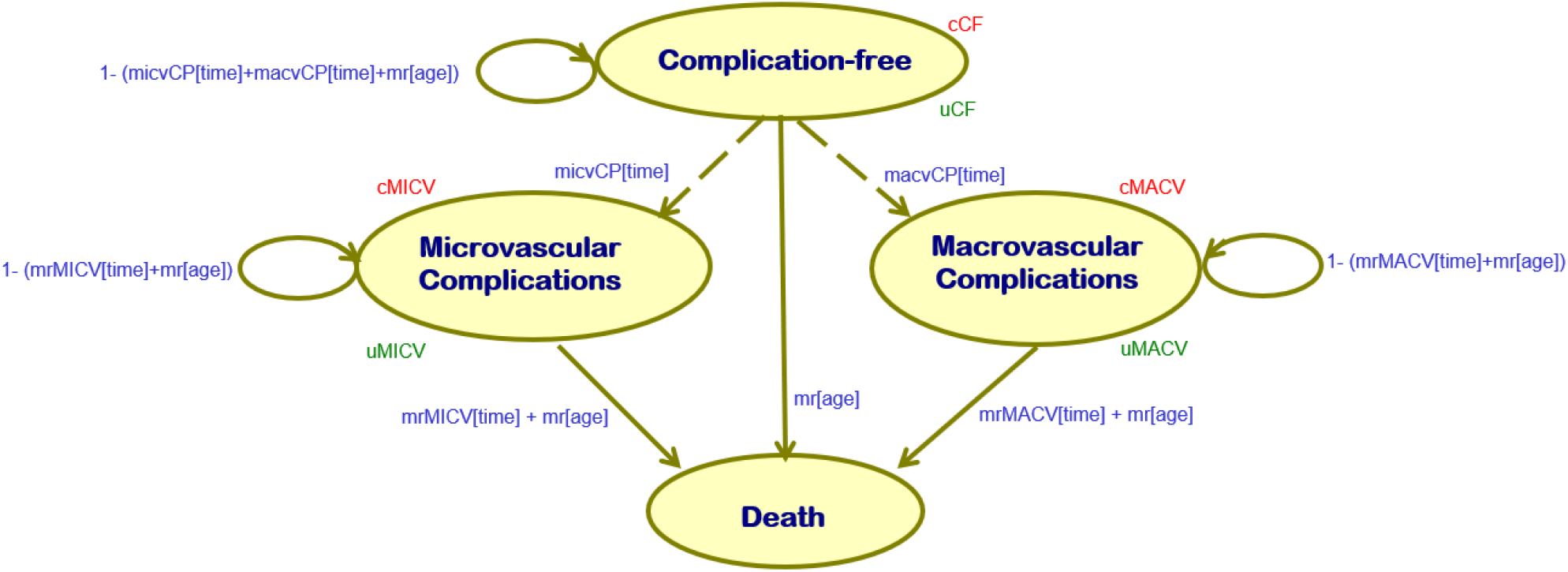
Diagrammatic representation of the Markov model

### Model parameters

From a survival analysis study done in a referral hospital in Ethiopia 524 adult type-2 diabetes patients were followed for a median of 7.4 years. The findings of the study estimated the probability of remaining free of any complications to be 0.76 at 5 years and 0.4 at 10 years. Similarly the probability of dying from a diabetes related complication was reported to be 0.02 and 0.039 at 5 and 10 years, respectively (11). Using this survival function at different time points, lambda (scale) and gamma(shape) parameters for a Weibull distribution were estimated after applying the following formula: **S(t)=exp (-λ t** ^**γ**^). (Table 2)

**Table 1.** Description of the interventions.

| Intervention | Comparator |
| --- | --- |
| FreeStyle Libre 2 | Standard Glucometer |
| A stand-alone continuous glucose monitoring device with a sensor that lasts for 2 weeks and does not require calibration with a standard glucometer. | A standard device used in usual practice to measure blood glucose level with a finger prick test. |

**Table 2.** Risk functions used in the Markov model.

| Change of Health State | Function | Parameter | Value (SE) | Distribution | Source |
| --- | --- | --- | --- | --- | --- |
| From complication-free to either of the complication states | Weibull | $\lambda$ | 0.0197<br>(0.000861) | Log Normal | (11) |
| | | $\gamma$ | 1.737<br>(0.0759) | | |
| From either of the complication states to death | | $\lambda_1$ | 0.00415<br>(0.00018) | Log Normal | (11) |
| | | $\gamma_1$ | 0.972<br>(0.0425) | | |

Costs were derived from a hospital-based study done in central Ethiopia that implemented a micro-costing technique for direct medical costs and the human capital approach for indirect costs (12). The reported costs, originally in 2020 US dollars, were adjusted to 2024 US dollars using the Consumer Price Index (CPI) inflation calculator and subsequently converted to Ethiopian Birr (ETB) for the model analysis (Appendix Table 6). Patients with microvascular complications were found to incur 1.6 times higher healthcare costs based on a systematic review (13), while expenses for patients with macrovascular complications were estimated to be twice as high, informed by expert opinion. The cost of the intervention was determined using the market price of the FreeStyle Libre device. For standard glucometer use, the cost estimation was based on an average self-monitoring frequency of three times per day.

A meta-analysis of EQ-5D based studies in Ethiopia reported the pooled utility value for patients with diabetes to be 0.78 (14). The additional disutility associated with microvascular and macrovascular complications is taken from another multi-center study in Ethiopia (15).

The DIAMOND randomized clinical trial measured the effects of CGM devices on glycemic control when compared to self-monitoring of blood glucose(SMBG) levels in 158 adults with type-1 diabetes (17). The total time spent outside the glycemic target range was 387 minutes/day in the control (SMBG) group and 243 minutes/day in the intervention (CGM) group. The relative risk reduction from using a continuous glucose monitoring device was 37.2%. This relative risk value was used in the Markov model to adjust the transition probabilities of developing diabetes-related complications and mortality.

Mortality from other causes is also considered in the model using Global Health Observatory’s age and gender specific life table for Ethiopia (16).

### Analysis

#### Base case

In the base case analysis, a deterministic approach was applied using the values provided in Table 3 as the model inputs. The analysis estimated the expected costs and health outcomes, measured in quality-adjusted life years (QALYs) gained, for both interventions. The results were summarized through the calculation of the incremental cost-effectiveness ratio (ICER). Additionally, the net monetary benefit was presented using a willingness-to-pay threshold set at one times the current GDP per capita for Ethiopia (18). To account for variations in demographic characteristics, ICER estimates (Table 5), and cost-effectiveness acceptability curves (CEACs) were generated for different age groups and sexes, informed by the life table for Ethiopia (Appendix Figure 5).

**Table 3.**
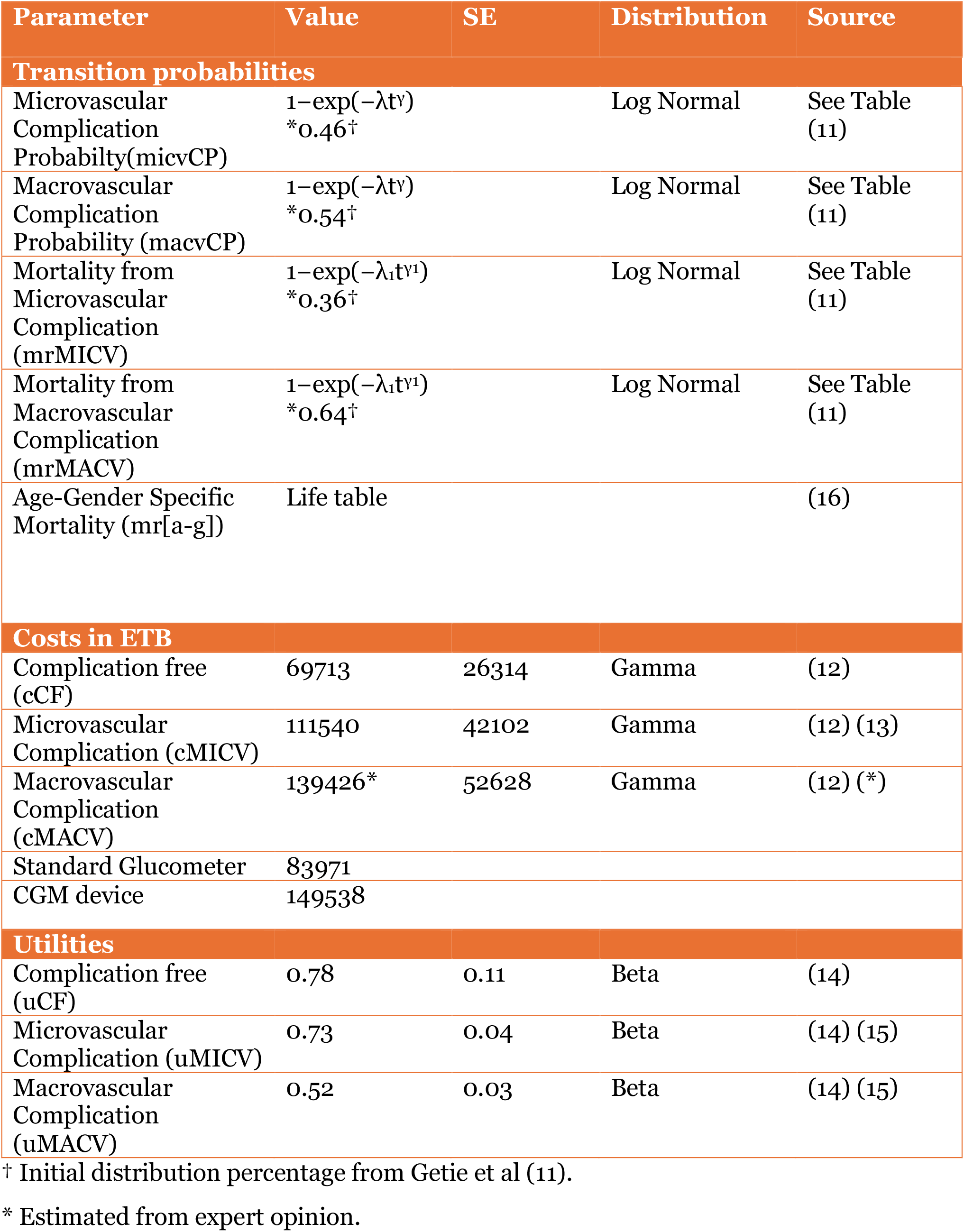
Yearly values of probabilities, cost and utility inputs used in the model.

**Table 4.**
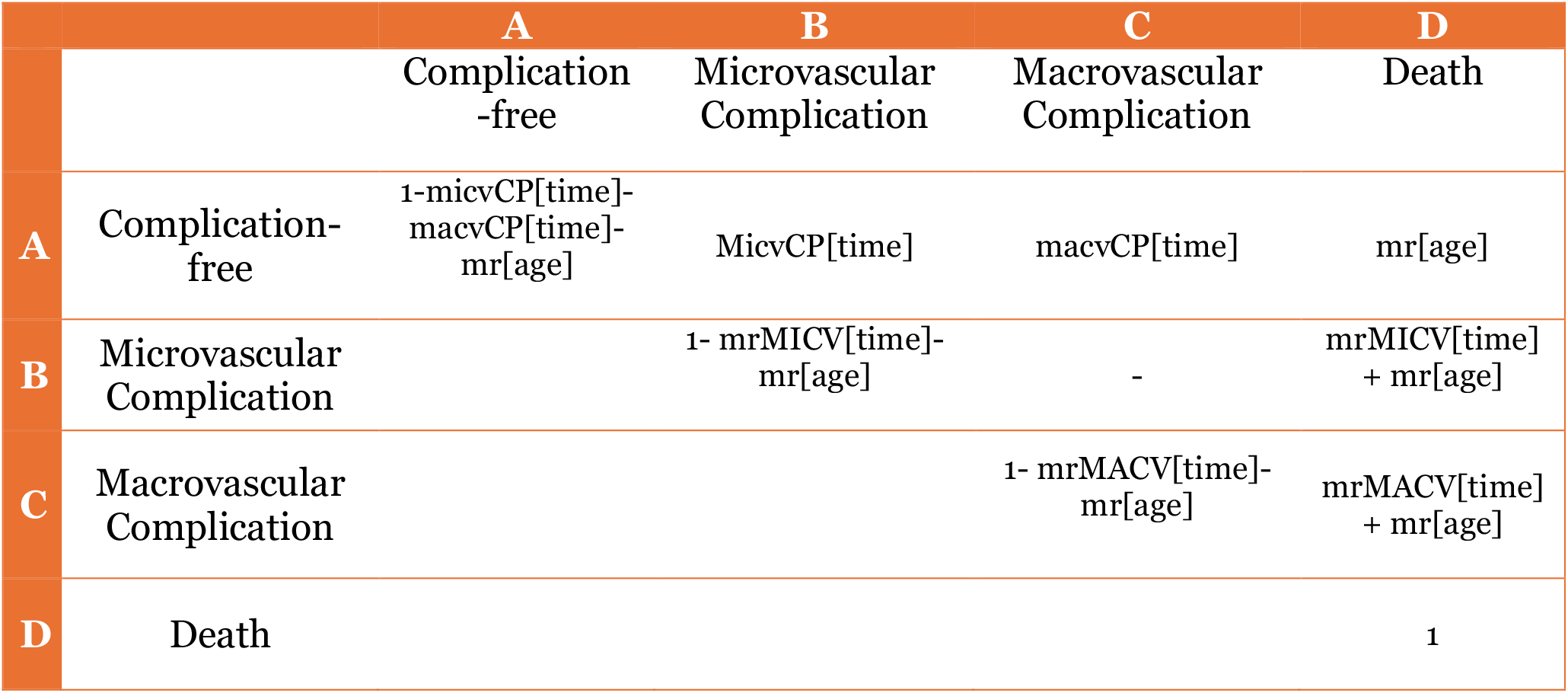
Transition probabilities matrix.

**Table 5.** Incremental cost effectiveness ratio and Net monetary benefit across age-gender variables==p(heterogenous sub-group analysis)

| Intervention | Cost | QALYs | Incremental Cost | Incremental QALYs | ICER | NMB<sup>†</sup> |
| --- | --- | --- | --- | --- | --- | --- |
| **Male, Age 20** |  |  |  |  |  |  |
| Standard | 1,999,892 | 11.37 |  |  |  |  |
| CGM | 2,208,956 | 12.71 | 209,064 | 1.34 | 155,625 | 12,036 |
| **Male, Age 40** |  |  |  |  |  |  |
| Standard | 1,782,696 | 10.22 |  |  |  |  |
| CGM | 1,906,679 | 11.13 | 123,983 | 0.92 | 135,353 | 16,447 |
| **Male, Age 60** |  |  |  |  |  |  |
| Standard | 1,255,414 | 7.45 |  |  |  |  |
| CGM | 1,279,430 | 7.85 | 24,016 | 0.40 | 59,718 | 6,282 |
| **Female, Age 20** |  |  |  |  |  |  |
| Standard | 2,018,460 | 11.47 |  |  |  |  |
| CGM | 2,236,407 | 12.86 | 217,947 | 1.39 | 156,881 | 72,469 |
| **Female, Age 40** |  |  |  |  |  |  |
| Standard | 1,833,700 | 10.48 |  |  |  |  |
| CGM | 1,972,224 | 11.47 | 138,524 | 0.99 | 140,290 | 23,060 |
| **Female, Age 60** |  |  |  |  |  |  |
| Standard | 1,344,966 | 7.93 |  |  |  |  |
| CGM | 1,376,239 | 8.38 | 31,273 | 0.45 | 69,791 | 4,459 |

<sup>†</sup> at a threshold level of one times GDP per capita in Ethiopia 2023 (ETB 165,000)(18)

A deterministic method may provide a reliable estimate of cost-effectiveness based on the best available data, but it does not consider the uncertainty of inputs to the model or the possibility of other clinical scenarios.

#### One-way sensitivity analysis

To test the robustness of the model’s conclusion to some of the assumptions, a one-way sensitivity analyses on all the costs, utilities and transition probabilities was performed and the results presented in a tornado diagram (Figure 2).

**Figure 2.**
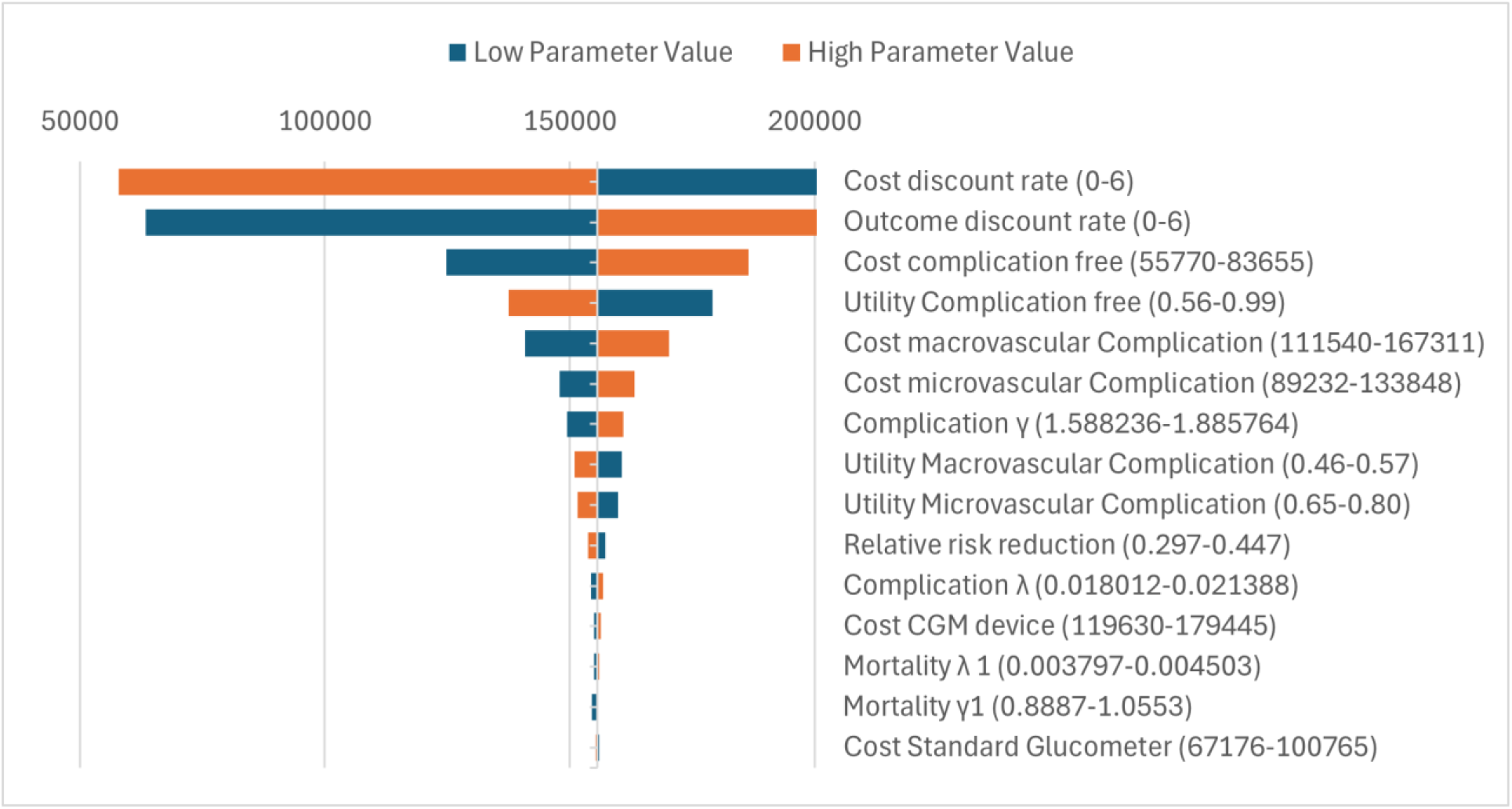
One-way sensitivity analysis for low and high parameter values

#### Probabilistic sensitivity analysis

Overall model uncertainty was analyzed with probabilistic sensitivity analyses (PSA), and the results are presented as cost-effectiveness acceptability curves, and scatterplots (Figure 3 and 4).

**Figure 3.**
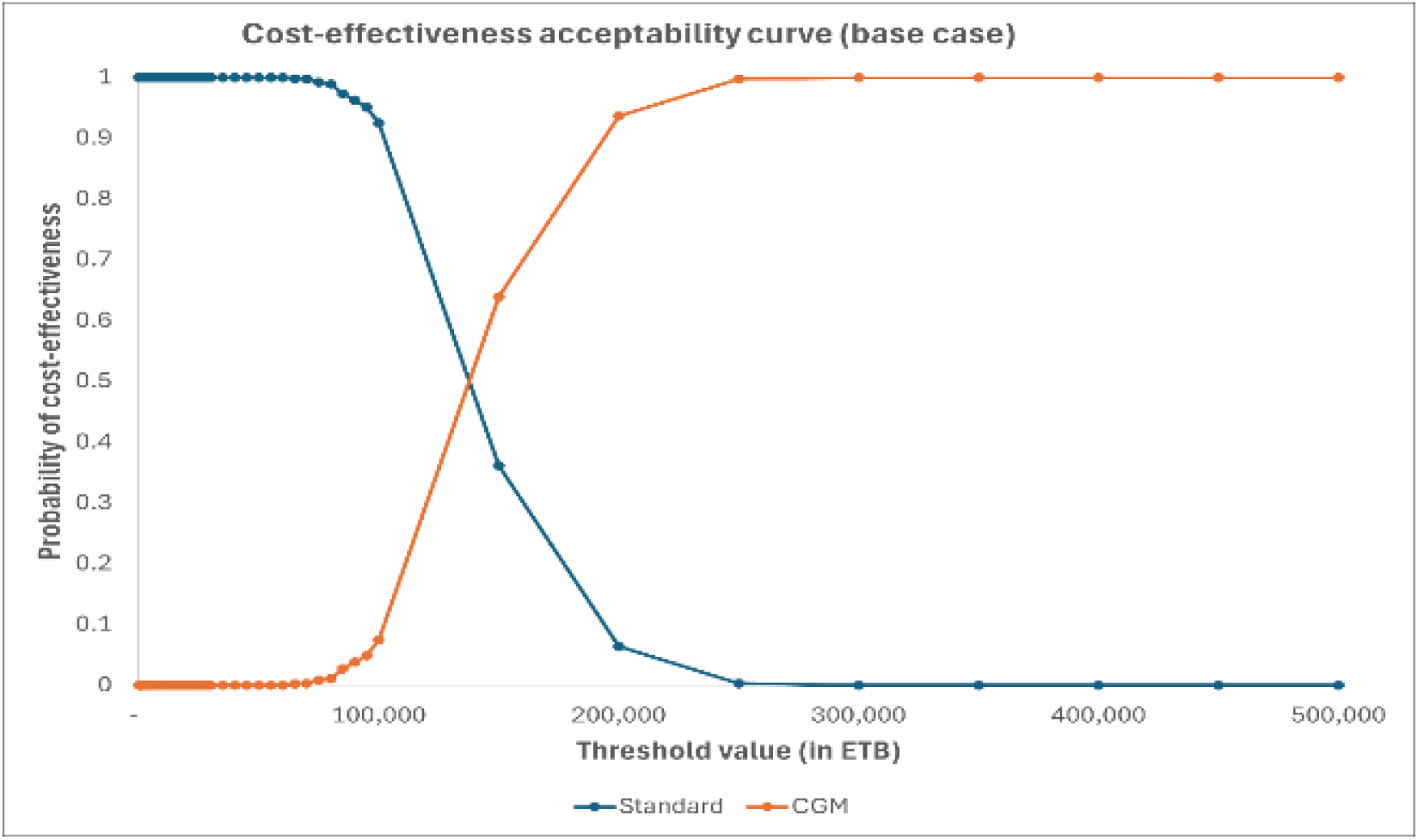
Cost-effectiveness acceptability curve for base case (female, age 20)

**Figure 4.**
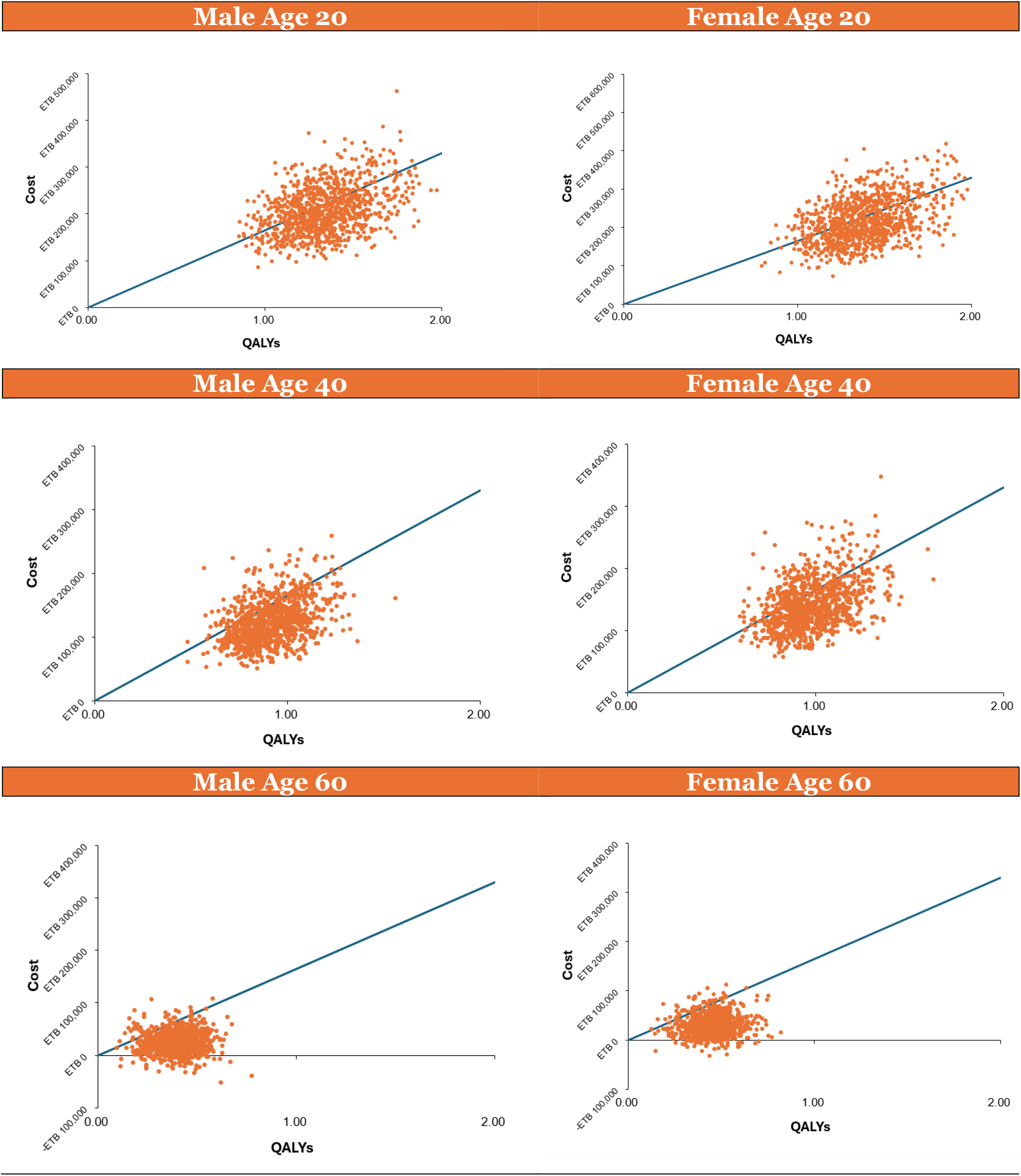
Cost effectiveness scatter plot with threshold level across age-gender variable

**Figure 5.**
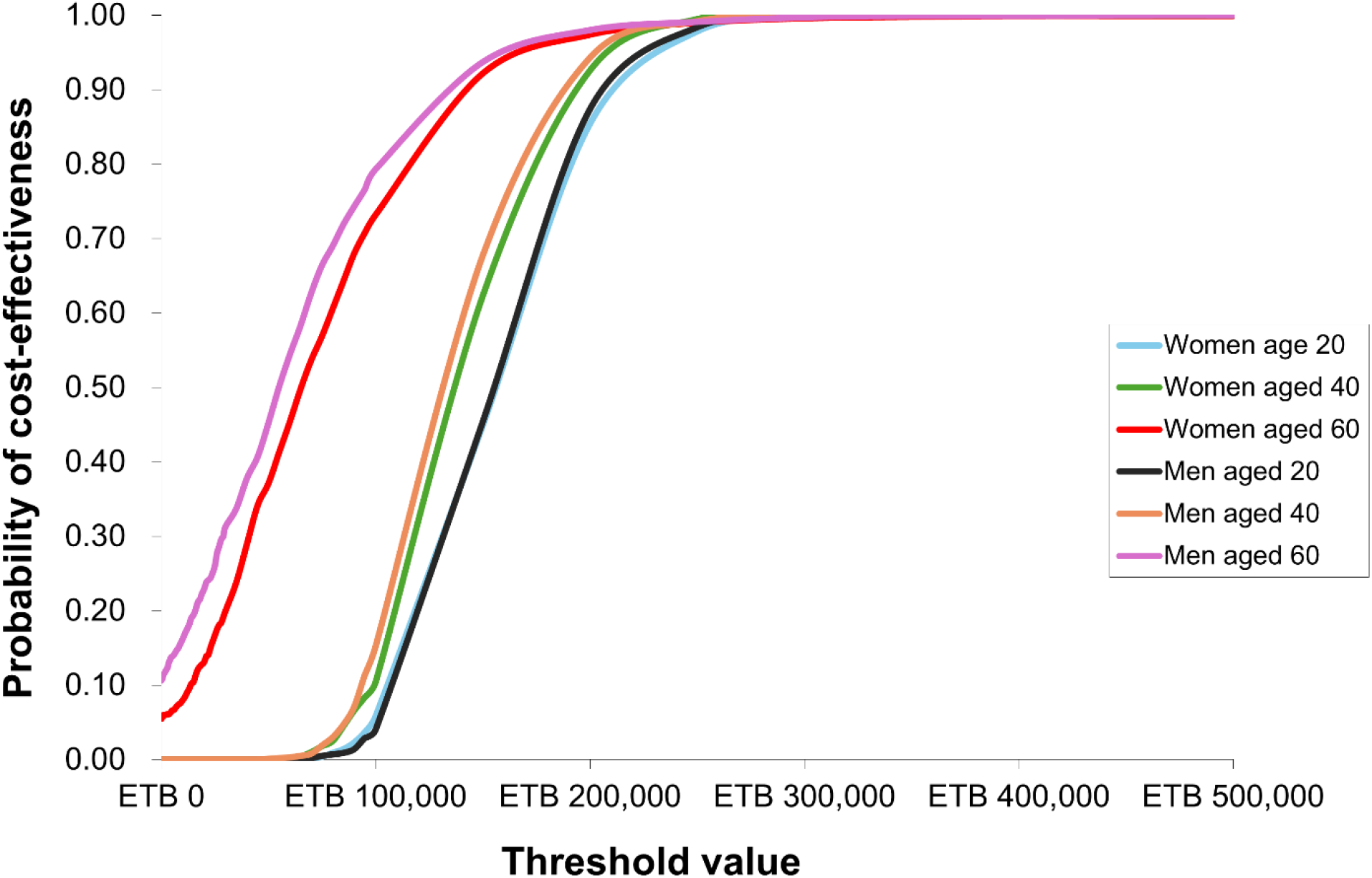
Cost-effectiveness acceptability curves across age-gender variables

A Markov chain Monte Carlo simulation with 1000 iterations was done using Excel v.2411. In the PSA, the variables in the model were replaced with distributions (See Table 3). We assumed the cost parameters to have a gamma distribution, the health outcome to follow a beta distribution and the transition probabilities to follow a log-normal distribution.

## Results

### Base case

The outcomes of this model are the incremental cost-effectiveness ratio (ICER) between the standard treatment and CGM measured as costs per QALYs gained and net monetary benefit (NMB) taking a threshold value of *1xGDP/capita* in Ethiopia (ETB 165,000). The calculations for this study are done with the sex parameters of male or female and the average age when entering the cohort of 20, 40 and 60 years. Table 5 summarizes the age- and sex-specific ICERs and NMB.

The incremental cost-effectiveness ratio (ICER) decreases with age in both sexes, indicating improved cost-effectiveness in older populations. However, females consistently exhibit a higher ICER than males, with the disparity between the sexes widening with age. A higher ICER reflects a lower “return on investment,” suggesting that greater financial resources are required to gain fewer QALYs, resulting in less favorable health outcomes.

### One-way sensitivity analysis

In the one-way sensitivity analysis, the highest ICER of ETB 156,881 per QALY (the case for a female starting cohort at age 40) is taken as the baseline conservatively.

Figure 2 presents a tornado diagram illustrating the sensitivity analysis of key parameters influencing the model. Each bar represents the impact of varying parameter values (low and high) on the model’s outcome, specifically the ICER. The ranges for each parameter are provided in parentheses, reflecting their plausible variability within the analysis.

The cost discount rate and outcome discount rate emerge as the most critical drivers, indicating that variations in these parameters can substantially alter the results of the model. Additionally, the costs associated with complication-free states and macrovascular complications are highly sensitive, suggesting that interventions aimed at reducing complications or improving complication-free periods could have significant cost implications. Utility parameters (e.g., for complication-free states and micro/macrovascular complications) also play a notable role, emphasizing the importance of quality-of-life adjustments in influencing model outcomes.

Parameters such as mortality rates, cost of continuous glucose monitoring (CGM) devices, and standard glucometer costs exhibit relatively minor impacts, indicating they are less pivotal in driving cost-effectiveness results within the modeled ranges. This suggests that while these factors are relevant, their variability does not fundamentally shift the model’s conclusions.

### Probabilistic sensitivity analysis

Figure 3 displays the cost-effectiveness acceptability curve (CEAC) for the highest ICER group in the base case analysis (female, age 20), illustrating the probability of cost-effectiveness for the two interventions; standard self-monitoring and continuous glucose monitoring (CGM)—across a range of threshold values in Ethiopian Birr (ETB).

The CEAC demonstrates the shift in cost-effectiveness between the two interventions as the threshold increases. At low threshold values (<100,000 ETB), standard care is the dominant option, with a near 100% probability of being cost-effective. However, as the threshold rises, the probability of CGM being cost-effective increases, overtaking standard care at approximately 150,000 ETB. Beyond this point, CGM consistently maintains a higher probability of cost-effectiveness, approaching certainty at thresholds exceeding 300,000 ETB (less than two times GDP/capita in Ethiopia).

This analysis indicates that CGM becomes the preferred intervention at higher threshold values, reflecting its greater cost-effectiveness for decision-makers willing to invest more in health outcomes. Conversely, at lower thresholds, standard care remains the more viable option, suggesting that its affordability plays a critical role in contexts with constrained resources.

### Sub-group analysis

Figure 4 shows a scatter plot of the distribution of ICER and QALY outcomes for males and females across different age groups relative to the threshold of ETB 165,000. Points below the threshold represent cost-effective outcomes.

#### Age 20 (Males and Females)

The majority of points lie near or above the threshold, indicating that the intervention is less likely to be cost-effective for younger individuals. The variability in QALY gains is higher for this age group, suggesting greater uncertainty in health benefits.

#### Age 40 (Males and Females)

There is a noticeable shift with more points closer to or below the threshold, reflecting improved cost-effectiveness compared to age 20. This suggests that middle-aged individuals may derive greater value from the intervention, making it more justifiable within the threshold.

#### Age 60 (Males and Females)

A significant portion of points falls below the threshold, indicating that the intervention becomes more cost-effective in older populations. This reflects the cumulative health benefits or reduced costs associated with managing chronic conditions in older individuals.

## Discussion

The results of this study illustrate the potential of continuous glucose monitoring (CGM) as a cost-effective intervention for diabetes management in Ethiopia. The base-case analysis reveals that the incremental cost-effectiveness ratio (ICER) varies significantly across age and gender groups. These findings suggest that the implementation of CGM is particularly favorable for older populations, slightly more for males, where the cumulative health benefits and reduced costs associated with diabetes-related complications justify the investment.

These findings align with previous global cost-effectiveness analyses of CGM, such as the DIAMOND randomized trial conducted in the United States, which also reported favorable ICERs for CGM use in insulin-dependent diabetes patients, particularly in older age groups (17). The DIAMOND study found an ICER of approximately $100,000 per QALY gained, with more pronounced benefits in populations with poor glycemic control, similar to the improved cost-effectiveness observed in older Ethiopian cohorts in this model.

A cost-effectiveness study conducted in Sweden similarly reported a decreasing ICER with age, as older patients often face higher baseline health costs and are more likely to benefit from enhanced glucose monitoring (19). However, both the DIAMOND trial and the Swedish analysis included high-income settings with greater healthcare resources, which may explain the significantly lower ICERs observed in Ethiopia due to baseline cost differences.

Notably, a health technology assessment in Ontario, Canada, reported higher ICERs for CGM use in adults with type 2 diabetes, suggesting cost-effectiveness only at higher willingness-to-pay thresholds (5). The difference may stem from the higher market prices of CGM devices in North America and greater reliance on healthcare subsidies compared to Ethiopia’s more out-of-pocket-based health financing system.

Within sub-Saharan Africa, limited economic evaluations have explored CGM’s impact. A systematic review on diabetes care costs in Ethiopia identified a significant financial burden associated with diabetes complications, emphasizing the potential value of preventive interventions (13).

The probabilistic sensitivity analysis (PSA) further confirms that CGM becomes the preferred intervention at GDP per capita thresholds exceeding ETB 150,000, with a near-certain probability of cost-effectiveness at thresholds above ETB 300,000. This demonstrates that while CGM is an economically viable option in scenarios with higher resource allocation, standard care remains dominant at lower thresholds, reflecting the affordability challenges in low-income settings like Ethiopia.

Sensitivity analyses also reveal that the most influential parameters are discount rates for costs and outcomes, as well as costs associated with complication-free and macrovascular states. Conversely, variations in CGM device costs or standard glucometer costs have a relatively minor impact on cost-effectiveness outcomes, indicating that the primary drivers are improvements in health outcomes and reductions in long-term complications.

The results highlight CGM’s potential to improve diabetes management by reducing the time patients spend outside target glucose ranges and lowering the risks of severe diabetes-related complications.

## Limitations

Despite the strengths of this study, several limitations must be acknowledged. The model simplifies the progression of diabetes into discrete states and may not fully capture the true complexity and variability of the disease’s natural course, including the simultaneous occurrence of micro and macro-vascular complications. The study uses data from Ethiopian hospitals and global literature, which may not fully represent all population groups or regional variations within Ethiopia. The cost analysis applies inflation-adjusted costs from previous studies, which might not account for current market fluctuations or unanticipated costs. In addition, non-financial barriers, such as lack of awareness, cultural acceptance, and the digital literacy required for CGM adoption, were not included in the analysis but may influence the feasibility of large-scale implementation.

### Generalizability

The findings are specific to Ethiopia’s healthcare context and may not be directly transferable to other low- and middle-income countries with differing healthcare systems and resource constraints.

## Conclusion

This study demonstrates that CGM is a cost-effective intervention for diabetes management in Ethiopia, particularly among older adults and males, when evaluated against a threshold of one times GDP per capita. While affordability remains a barrier for younger and economically disadvantaged populations, the potential health benefits and reduced complications make CGM a promising alternative to standard care. The findings emphasize the importance of age- and gender-specific strategies in the implementation of CGM technologies to maximize health outcomes and economic efficiency.

Future research should focus on refining parameter estimates, reducing model uncertainty, incorporating real-world data on CGM adoption, and exploring different financing mechanisms to enhance affordability and accessibility. Policymakers should consider these findings to develop targeted interventions that address the unique needs of diverse patient groups, ultimately contributing to improved diabetes care and reduced healthcare burden in Ethiopia.

## Data Availability

All data produced in the present work are contained in the manuscript

## Appendix

**Table 6.** Direct and indirect costs for diabetes patients in Ethiopian Birr (ETB)

| Cost items | Average yearly cost (ETB) |
| --- | --- |
| Direct medical cost | **25213.56** |
| Cost of registration/ folder | **1561.90** |
| Cost of consultation fees | **1692.06** |
| Cost of Insulin | **17069.36** |
| Cost of Insulin syringe/ disposable | **2231.29** |
| Cost of laboratory investigations | **2658.95** |
| Direct non-medical cost | **22440.66** |
| Cost of non-prescription items | **3588.32** |
| Cost of dietary modification | **8236.41** |
| Cost of transportation | **10616.23** |
| Total Indirect cost | **22059.04** |
| Patients' income lost (ETB) | **10846.98** |
| Patients absent from work (in days) | 14.52 |
| Patients travel time to the hospital (in minute) | 460.2 |
| Accompany income lost (ETB) | **11214.06** |
| Accompany absent from work (in days) | 15.24 |
| Accompany travel time to hospital (in minute) | 433.08 |
| Patients and accompany absent from work (in days) | 25.68 |
| Total cost of diabetes | **69713.26** |

*\*taken from a study in hospitals in Southwest Shewa zone, Central Ethiopia reported as 2020 USD, adjusted for inflation through CPI to 2024 USD and converted to ETB (12)*

